# Advanced bedside monitoring for patients with acute respiratory failure during non-invasive respiratory support: A systematic review and meta-analysis

**DOI:** 10.64898/2026.02.05.26345575

**Authors:** Brigitta Fazzini, Timothy J Stephens, Florence Pickles, Georgia Mathieson, Richard Pattison, Eileen Kelly, Saira Nazeer, Leo Heunks, Jonne Doorduin, Zudin Puthucheary

**Affiliations:** Critical Care and Perioperative Medicine Research Group, William Harvey Research Institute, Barts and The London School of Medicine and Dentistry, Queen Mary University of London, London, UK; Adult Critical Care Unit, Royal London Hospital, Barts Health NHS Trust, London, UK; Department of Speech and Language Therapy, Royal London Hospital, Barts Health NHS Trust; Department of Critical Care Medicine, Radboud University Nijmegen Medical Centre, Netherlands

**Keywords:** advanced monitoring, respiratory muscle workload, respiratory effort, non-invasive respiratory support, acute respiratory failure

## Abstract

**Background:** Patients with acute respiratory failure requiring non-invasive respiratory support are at high risk of deterioration. Different advanced monitoring instruments are available that can provide objective measurements. However, there is currently no evidence synthesis on these instruments. The aim of this project is to systematically synthetise data identifying the advanced monitoring instruments used and their effectiveness.

**Methods:** We conducted a systematic search of MEDLINE (via Pubmed), EMBASE, Web of Science, Cochrane Library and CINAHL (PROSPERO registration: CRD42024597047). We included studies with acute respiratory failure patients requiring non-invasive respiratory support where the investigators used advanced monitoring instrument during hospital stay. We followed the Preferred Reporting Items for Systematic Reviews and Meta-Analyses (PRISMA) guidelines.

**Results:** Seventy-eight studies including 3709 patients fulfilled the selection criteria. The monitoring instruments used were respiratory muscle ultrasound in 32% (n= 25/78), oesophageal manometry in 32% (n= 25/78), electrical impedance tomography in 24% (n= 19/78), electrical activity of the diaphragm (Eadi) catheter in 18% (n= 14/78) and surface EMG of parasternal muscle in 6% (n= 5/78). Thirteen studies (17%) used a multi-modal monitoring approach. Patients failing non-invasive respiratory support showed higher oesophageal pressure (ΔPes) [MD 12.60 (95% CI 4.03;21.16), p=0.004], lung ultrasound score (LUS) [MD 3.93 (95% CI 1.29,6.570), p=0.003] and parasternal intercostal thickening fraction (PIC-TF%) [MD 12.58 (95% CI 8.02,17.13), p<0.001] but lower diaphragmatic thickening fraction (DTF%) [MD - 17.20 (95% CI −20.97,-13.42); p<0.001] and lower diaphragmatic excursion (DE) [MD - 0.95 (95% CI −1.08,-0.82); p<0.001.

**Conclusion:** Advanced monitoring instruments may detect patient failing non-invasive respiratory support.

**Take home message:** Advanced bedside monitoring during non-invasive respiratory support can provide unique physiological insights into respiratory muscle workload and treatment response in acute respiratory failure. Our meta-analysis shows that five measurements: i) oesophageal pressure changes (ΔPes), ii) lung ultrasound score (LUS), iii) parasternal intercostal thickening fraction (PIC-TF%), iv) diaphragmatic thickening fraction (DTF%) and v) diaphragmatic excursion (DE) may discriminate patients who are responders from non-responders to non-invasive respiratory support.

## Introduction

Acute respiratory failure (ARF) is one of the leading causes of hospital admission for more than 600,000 patients in the UK and accounts for 2.6 million deaths worldwide (1–3). Non-invasive respiratory support is increasingly used across different hospital settings as a first line treatment for the management of acute respiratory failure (4,5). Modalities such as high-flow nasal oxygen (HFNO), continuous positive airway pressure (CPAP), and bilevel positive airway pressure (BiPAP), offer the advantage of being readily accessible and applicable to all patients, even outside critical care. However, patients requiring non-invasive respiratory support remain at high risk of treatment failure (defined as intubation or death), which is often recognised late, largely because traditional monitoring relies on delayed or indirect indicators of respiratory failure (6,7). These system-level failures highlight the importance for improved physiological assessment instruments that can support more timely clinical decision-making.

Traditional bedside monitoring such as vital signs and composite scores like respiratory rate-oxygenation (ROX) index (8),the heart rate, acidosis, consciousness, oxygenation, and respiratory rate (HACOR) score (9) and early warning scores (NEWS2) (10) can identify patients at high risk of deterioration. However, these only reflect the downstream consequences of respiratory imbalance (such as hypoxaemia or tachypnoea) rather than the physiological mechanism underlying respiratory distress (11–16). In contrast, monitoring the physiological balance between the respiratory load and the respiratory muscle capacity provides a window to detect failure at an early stage (17–20). Without such mechanistic insight individualized titration of treatment support is cumbersome.

Several advanced bedside instruments have been developed to quantify different domains of the respiratory balance including respiratory effort, neural respiratory drive, respiratory muscle efficiency, and lung aeration or ventilation distribution directly or through proxy measures (21–27). These advanced instruments may provide physiological indicators of respiratory failure progression and treatment failure potentially detecting maladaptive responses in an early phase (16,28–30). Despite many studies demonstrate improvements in different physiological parameters (e.g. reduced inspiratory effort or enhanced lung aeration), it remains unclear whether these changes consistently translate into better clinical outcomes. Existing studies also use different measurements making it difficult to determine which instruments or parameters are clinically meaningful and limit their adoption. This uncertainty highlights a major gap in current evidence and the need for a systematic synthesis to identify relevant instruments, describe the physiological domains and parameters measured, and evaluate their clinical utility in detecting treatment failure for guiding decision-making.

To address these gaps, this systematic review and meta-analysis therefore aim to:

1. identify and describe advanced bedside monitoring instruments used to assess the respiratory load-capacity balance and lung aeration during non-invasive respiratory support.
2. summarize the physiological measurements obtained from each monitoring instrument and examine how these vary across different non-invasive respiratory support modalities.
3. evaluate the effectiveness of these monitoring instruments in detecting treatment failure during non-invasive respiratory support.

## Methods

The study protocol for this review was registered with the PROSPERO online database of systematic reviews (CRD42024597047). We conducted this systematic review which include a meta-analysis and meta-synthesis following the PRISMA guidelines, and the Cochrane Handbook for systematic reviews (31,32). PRISMA checklist is available in supplementary file (Table S4).

### Definitions

The targeted condition was defined as acute hypoxaemic respiratory failure with hypoxia (i.e. arterial oxygen tension (P_a_O_2_) of <8.0 kPa) and with or without hypercapnia (i.e. arterial carbon dioxide tension (P_a_CO_2_) of >6.0 kPa). Acute respiratory failure is heterogenous with multiple overlapping causes including acute pneumonia on chronic obstructive pulmonary disease (COPD). Therefore, we accepted all various causes leading to acute respiratory failure, including acute on chronic conditions.

Non-invasive respiratory support were defined as all modalities supporting spontaneous breathing by delivering pressurised oxygen and support by flow or positive pressure through nasal prongs, oronasal, face-mask or helmet interface and without insertion of an invasive airways (such as endotracheal tube or tracheostomy) (33). These non-invasive respiratory support devices include high-flow nasal oxygen (HFNO), continuous positive airway pressure (CPAP) and bilevel positive airway pressure (BiPAP) (also referred to as non-invasive ventilation).

Advanced respiratory monitoring is an umbrella term used by experts to define any instrument able to assess the respiratory load and respiratory muscle capacity either by direct or proxy measurements (21,22).

Two expert consensus defined physiological measurements as the quantitative variables (acquired with the advanced instruments) that describe or derive from monitoring respiratory muscle function, including both direct measures of physiological processes and calculated indices derived from them (21,22). These encompass measurements of respiratory effort, neural drive, ventilation distribution, and lung ventilation. Derived indices that integrate or interpret these raw signals such as patient–ventilator asynchrony indices, neuromechanical coupling ratios, or pressure–time products were also included within this definition, as they reflect the interaction and performance of physiological systems under support (21,22). Therefore, in this review, physiological measurements are intended as any directly measured or computationally derived quantitative variable that characterises the respiratory load and respiratory muscle capacity (i.e. mechanics, muscle workload, neural drive) or ventilatory pattern during non-invasive respiratory support.

We defined treatment “failure” when patients did not improve and were either intubated and/or died while treated with non-invasive respiratory support in hospital. Conversely, we defined “success” when patients improved while on non-invasive respiratory support.

### Search strategy

We conducted a search across the following databases MEDLINE (via Pubmed), EMBASE, Web of Science, Cochrane Library and CINAHL. Systematic data search included the following Mesh terms/key words and Boolean operators (full searches is available in the supplementary materials). Below is an example showing the Medline search:

#1.

((acute respiratory failure) OR (ARDS) OR (acute respiratory distress syndrome))

#2 AND

((high-flow nasal oxygen) OR (continuous positive airway pressure) OR (CPAP) OR (bilevel positive airway pressure) OR (B*PAP) OR (non*invasive ventilation) OR (NIV))

#3 AND

((respiratory monitoring) OR (clinical evaluation) OR (respiratory rate) OR (heart rate) OR (tachypnoea) OR (accessory muscle activity) OR (agitation) OR (work of breathing) OR (WOB) OR (lack of cooperation) OR (outcomes predictors) OR (arterial blood gas) OR (ABG) OR (gas exchange) OR (pulse oximetry) OR (hypoxia) OR (expired CO2) OR (PCO2) OR (blood lactate levels) OR (transcutaneous CO2 monitoring) OR (TOSCA) OR (chest radiography) OR (chest x-ray) OR (CXR) OR (ultrasonography) OR (US) OR (respiratory muscle and chest ultrasonography) OR (electromyogra*) OR (EMG) OR (electrical impedance tomography) OR (EIT) OR (electrical impedance tomography) OR (EIT) OR (*esophageal manometry) OR (pleural pressure) OR (*esophageal pressure) OR (chest computed tomography) OR (CT chest) OR (CT))

We were expecting that studies may include different basic and advanced monitoring (i.e. respiratory rate and oesophageal manometry) or imaging as comparator. Therefore, the search was purposely wide to be able to capture all studies including all type of advanced respiratory monitoring to avoid missing records.

Studies were also identified and retrieved by citation searching from the references of each relevant study. References list and published systematic reviews were searched and screened to reduce the potential risk of missing data. The bibliographies of those papers that matched the eligibility criteria described above were also searched by hand to identify any further, relevant references, which were subject to the same screening and selection process. Five investigators (BF, FP, GM, RP, SN) independently screened title and abstracts in duplicate for selection of full-text review using Covidence Software (Covidence systematic review software, Veritas Health Innovation, Melbourne, Australia. Available at Link: www.covidence.org.) due to the high volume of records retrieved. If a decision was not achieved from reading the title and abstract alone, the full text was reviewed. The reviewers also independently reviewed the full text of relevant studies and decided on eligibility. Inter-rater disagreements in the study selection were resolved by consensus or if necessary, by consultation with a senior author (TS).

#### Inclusion and exclusion criteria

We included all studies including acute respiratory failure patients supported on the above mentioned non-invasive respiratory support and assessed/monitored by at least one advanced monitoring instrument. Eligible studies included adult patients (age > 18 years old) independent of sex/age/ethnicity admitted to any hospital ward with acute respiratory failure undergoing non-invasive respiratory support (i.e. HFNO, CPAP, BiPAP) and received assessment with advanced monitoring. Large treatment effect has been reported in studies including fewer patients (34), hence, we included all peer-reviewed studies comprising leastwise 10 patients as the minimum acceptable sample size recommended for physiological studies (35). We excluded studies published prior January 1^st^ 2000 as this is the date of the first landmark studies using non-invasive respiratory support (i.e. CPAP) was first published on the year 2000 (36). We also excluded: i) studies including patients receiving non-invasive respiratory support for other reasons than acute respiratory failure i.e. for palliative care/end-of life treatment comfort care, as preventive measure post-extubation, or via tracheostomy as long-term ventilation mode; ii) commentaries, editorials, reviews, conference abstract, posters, qualitative studies, guidelines and consensus report, protocol studies, iii) studies not peer-reviewed. A flow-chart of the whole process is presented according to PRISMA guidelines in Figure 2.

#### Data extraction

Five reviewers (BF, FP, GM, RP, SN) extracted the following information from each publication: author and year of publication, country, study design, setting, number of included patients, type of population, advanced monitoring instruments, specific measurements detected, physiological/clinical outcomes measured. Data extraction was performed in duplicate by four authors acting independently (BF, FP, GM, RP, SN). Authors were contacted by email if necessary or if data were missing. If no response, two reminders were sent, or other authors of the team were contacted. The characteristics of the studies included are presented in Table 1 of the supplementary materials.

**Table 1.** Summary of the clinical application of each advanced monitoring instrument: what they measure and how have been used in all 78 included studies.

| Advanced monitoring instruments | Physiological signals | Physiological variables | Domain measured |
| --- | --- | --- | --- |
| **Oesophageal manometry**<br>(17–19,29,48,49,52,53,56,62,67,69,70,72,74,77,106) | Oesophageal pressure (Pes)<br>Transpulmonary pressure (PL = Paw – Pes) | Oesophageal pressure swing (ΔPes)<br>Transdiaphragmatic pressure swing (Pdi/ΔPdi)<br>Pressure–time products (PTPdi/PTPes)<br>Transpulmonary pressure swing (ΔPI) | Respiratory effort<br>Neural respiratory drive<br>Lung distending pressure<br>Inspiratory load |
| **Electrical Impedance Tomography**<br>(EIT)<br>(101,41,102,103,43,51,54,57,62,106,60,61,63,65,66,69,73,78,70) | Thoracic impedance | Estimated tidal volume<br>End expiratory lung impedance (EELI/ΔEELI)<br>Global Inhomogeneity index (GI index)<br>Center of Ventilation (CoV%) | Respiratory load (global and regional ventilation distribution and ventilation-perfusion matching) |
| **Eadi catheter/oesophageal diaphragmatic EMG**<br>(42,44,45,104,50,90,55,58,95,64,107,68,108,109) | Electrical activity of the diaphragm (Eadi) | Electrical activity of the diaphragm signal (ΔEadi)<br>Diaphragmatic neuromuscular coupling<br>Transdiaphragmatic pressure/change in diaphragm electrical activity (ΔPdi/ΔEadi)<br>Asynchrony index | Neural respiratory drive (magnitude and timing)<br>Neuromechanical coupling |
| **Surface Electromyography**<br>(EMG)<br>(30,47,71,75,76) | Electrical activity of parasternal and abdominal muscles | Parasternal intercostal electromyography signal ($EMG_{para}/\Delta EMG_{para}$)<br>Diaphragmatic muscle electromyography signal ($EMG_{diaph}/\Delta EMG_{diaph}$)<br>Abdominal muscle electromyography signal ($EMG_{abdo}/\Delta EMG_{abdo}$)<br>Asynchrony index | Neural respiratory drive (magnitude and timing)<br>Extradiaphragmatic neural respiratory drive (magnitude and timing) |
| **Ultrasonography**<br>(US)<br>(80,81,79,82–86,46,87–89,91,93,112,105,57,94,59,96,28,97–100,113) | Diaphragm and parasternal intercostal muscle ultrasound images<br>Lung ultrasound images | Parasternal thickness/thickening fraction<br>Diaphragmatic thickness/thickening fraction<br>Diaphragmatic excursion<br>Lung ultrasound score | Diaphragm and parasternal intercostal muscle function (thickness and excursion)<br>Lung aeration |

#### Quality assessment

Two reviewers (BF, FP) independently assessed the risk of bias using the Newcastle-Ottawa Scale (NOS) for observational studies (31,32,37) and the Cochrane Risk of Bias tool 2 (ROB2) was used to for assessing randomized controlled trial (38). In case of any conflicts these were resolved following discussion with a senior reviewer (TS).

#### Data synthesis and analysis

A narrative and tabular synthesis of the findings were provided. Numerical data was collected for quantitative analysis.

Mean and standard deviation (SD) or median and interquartile range (1^st^ quartile to 3^rd^ quartile) were used for numerical data if appropriate, while odds ratio (OR) with 95% confidence interval (CI) were used for categorical data. For data presenting median and inter-quartile range (IQR) or median and range, mean and standard deviation (SD) were transformed according to standard equations (38). The studies included for meta-analysis were pooled together using the random-effects model accounting for the incidence. The results are presented in forest plots. Heterogeneity among studies was evaluated using the Tau^2^ test, I^2^ statistics and Cochrane Q. A two-sided p value ≤ 0.05 indicated statistical significance (38–40). Analysis of data was performed using the statistical software package RevMan 5.4.1^®^ (Review Manager, RevMan, [Computer program] Version 5.4, Copenhagen: The Nordic Cochrane Centre, The Cochrane Collaboration, 2020).

## Results

We identified 11710 studies through our literature search. After removing duplicates and publications that did not match our inclusion criteria, we were left with 78 publications. Figure 1 illustrates the flow diagram of the studies included.

**Figure 1.**
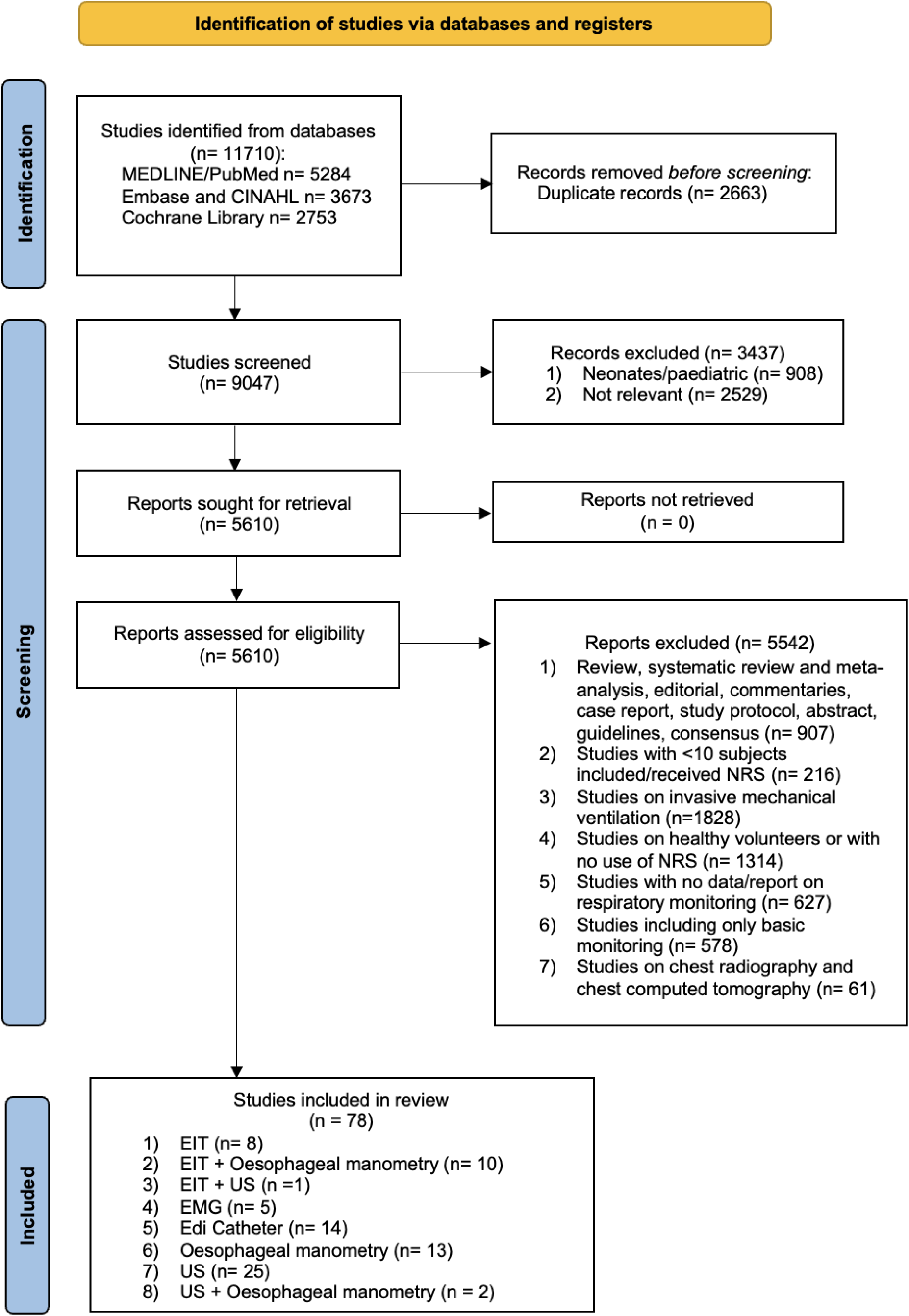
Flow diagram of selected studies according to the PRISMA guidelines.

Overall, the publications reported data on 3709 patients with acute respiratory failure requiring non-invasive respiratory support managed across different hospital settings including the emergency department, wards, high-dependency unit (HDU) and ICU. We found seventy single centre and six multicentre studies across Europe, Asia, North and South America. Studies characteristics are summarised in Table 1 of the supplementary materials (Table S1).

### Study quality appraisal

The Newcastle-Ottawa Scale found 51 studies of high quality (7-8*) (41–71,19,29,72,18,17,73–78,30) and 17 studies with medium risk of bias (5-6*) (79–96,28,97–100). The ROB2 found that the 10 randomised trials (101–105,94,106–109) had good quality with some overall concerns regarding bias related to the nature of the interventions (i.e. non-invasive respiratory support) and monitoring instruments which cannot be blinded. See Table 2 and table 3 of the supplementary materials (Table S2 and Table S3).

**Table 2.**
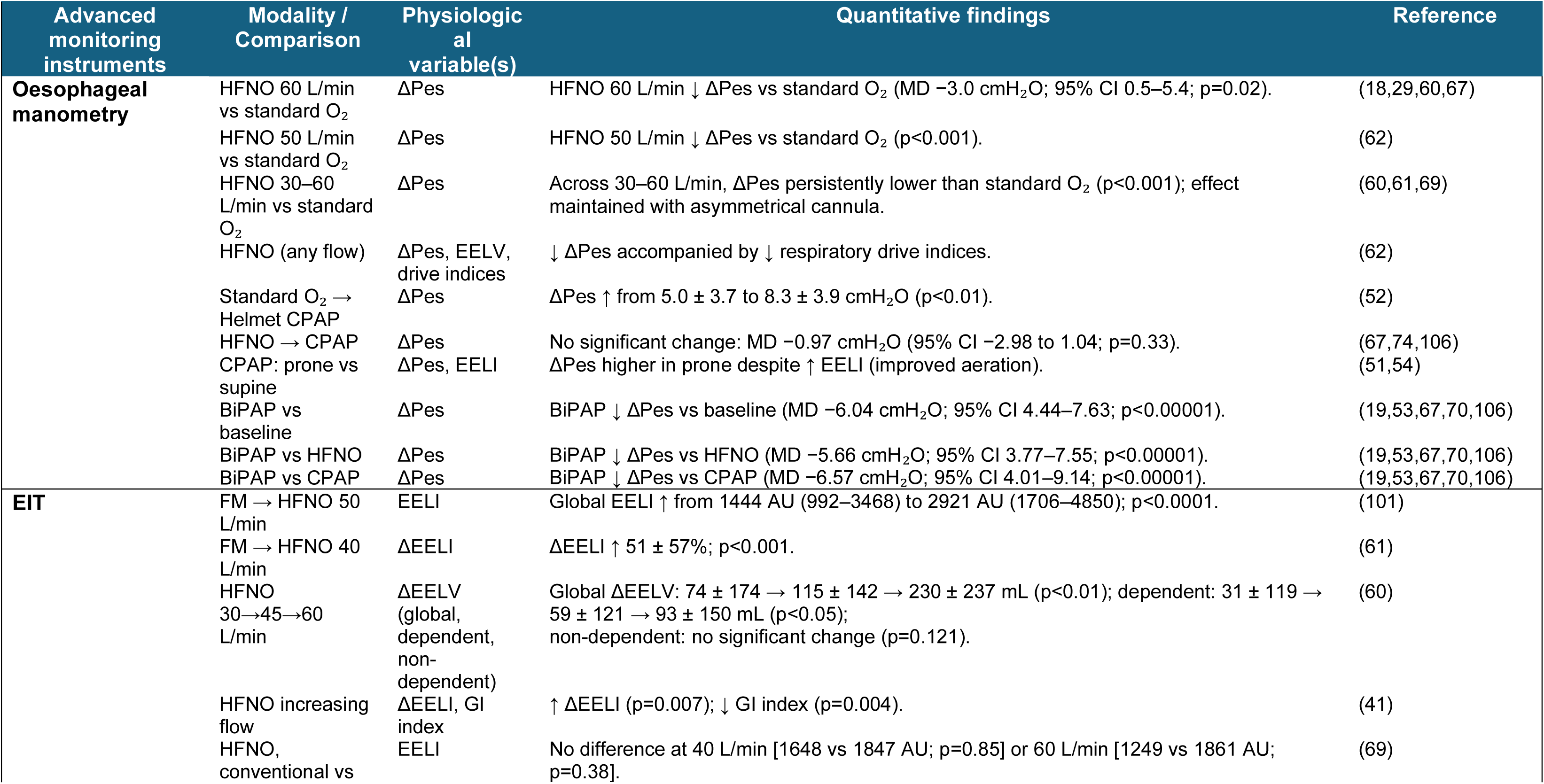
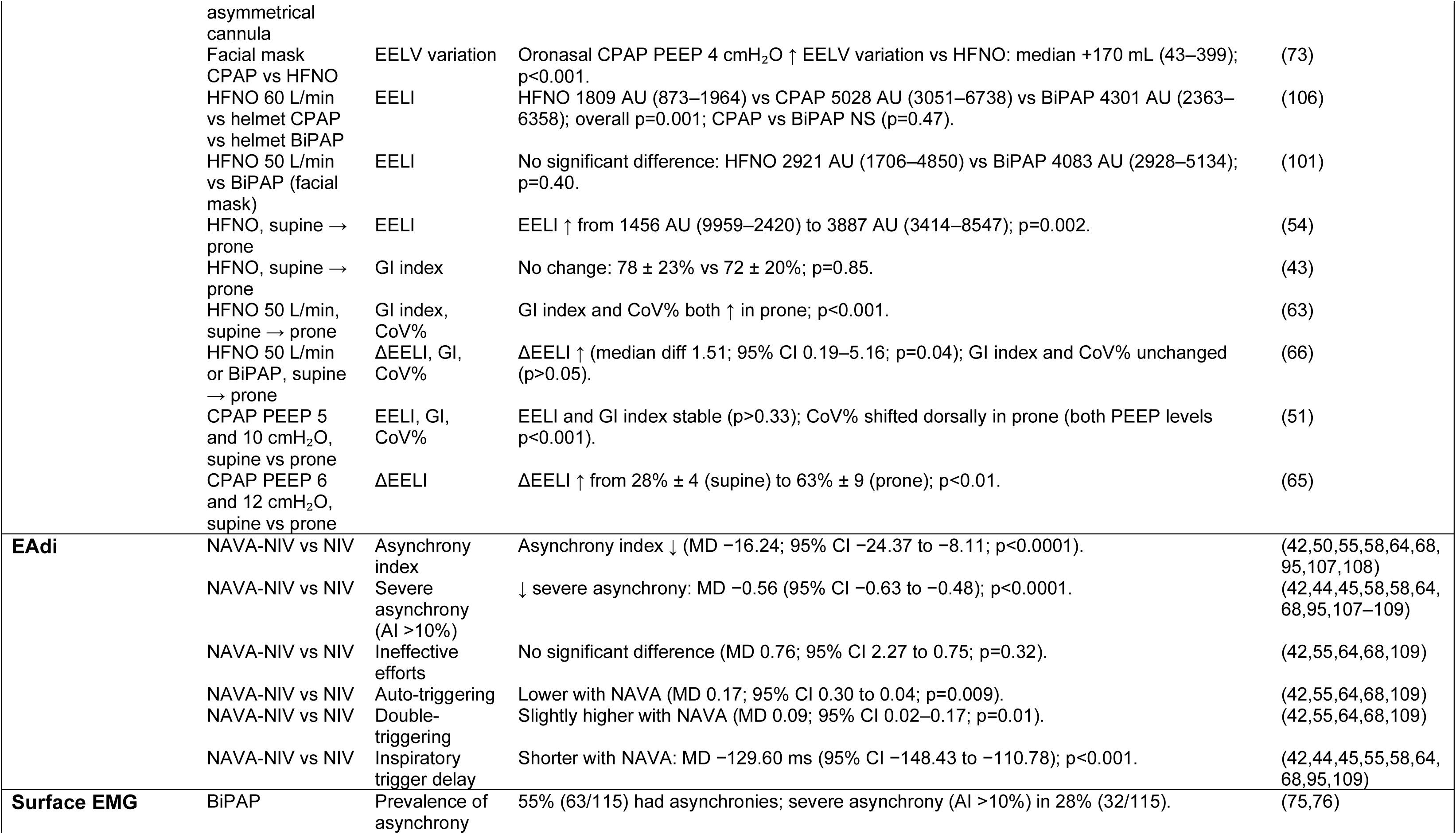
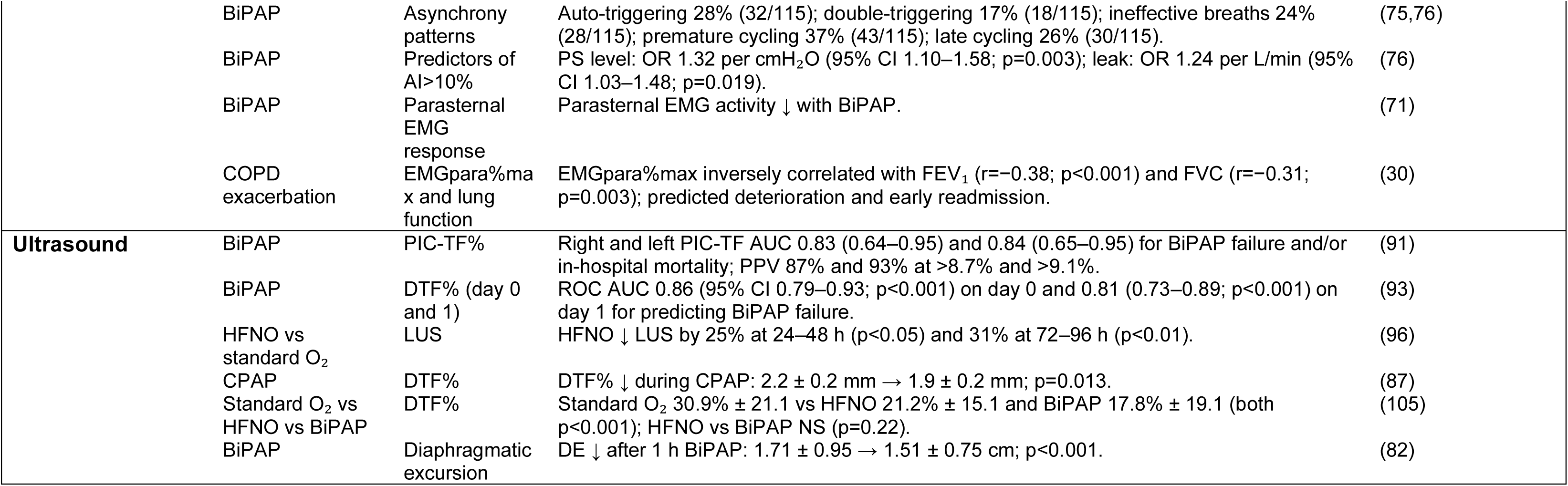
Physiological response during non-invasive respiratory support measured by advanced monitoring instruments. Abbreviations: AI: asynchrony index; AU: arbitrary units; BiPAP: bilevel positive airway pressure; CoV%: centre of ventilation; CPAP: continuous positive airway pressure; DE: diaphragmatic excursion; DTF: diaphragmatic thickening fraction; ΔDTF: change in diaphragmatic thickening fraction; EAdi: electrical activity of the diaphragm; EELI: end-expiratory lung impedance; ΔEELI: change in end-expiratory lung impedance; EELV: end-expiratory lung volume; ΔEELV: change in end-expiratory lung volume; EIT: electrical impedance tomography; EMG: electromyography; EMGpara: parasternal electromyography; FM: face-mask oxygen; GI index: global inhomogeneity index; HFNO: high-flow nasal oxygen; LUS: lung ultrasound score; MD: mean difference; NAVA: neurally adjusted ventilatory assist; NIV: non-invasive ventilation; NRS: non-invasive respiratory support; OR: odds ratio; Pes: oesophageal pressure; ΔPes: oesophageal pressure swing; PIC: parasternal intercostal muscle; PIC-TF: parasternal intercostal thickening fraction; PEEP: positive end-expiratory pressure; PS: pressure support; ROC: receiver operating characteristic; US: ultrasound; Paw: airway pressure; Pdi: transdiaphragmatic pressure; ΔPdi: transdiaphragmatic pressure swing; PTP: pressure–time product.

#### 1. Advanced monitoring instruments available at bedside

Of 78 studies, 24% (n= 19/78) used electrical impedance tomography (EIT) (101,41,102,103,43,51,54,57,62,106,60,61,63,65,66,69,73,78,70), 6% (n= 5/78) used surface EMG of parasternal muscle (30,47,71,75,76), 18% (n= 14/78) used electrical activity of the diaphragm (Eadi) (42,44,45,104,50,90,55,58,95,64,107,68,108,109), 32% (n= 25/78) used oesophageal manometry (17–19,28,29,48,49,52–54,56,60–62,67,69,70,72,74,77,102,106,110,111), and 32% (n= 25/78) used respiratory muscle ultrasound (80,81,79,82–86,46,87–89,91,93,112,105,57,94,59,96,28,97–100,113,111). Amongst these, thirteen studies (17%) employed a multi-modal monitoring approach; of which 13% (n= 10/78) combined EIT and oesophageal manometry, 3% (n= 2/78) used EIT and respiratory muscle ultrasound, and 3% (n= 2/78) used oesophageal manometry and US. (See Figure 1 and Table S1).

#### 2. Physiological measurements and response during different non-invasive respiratory support

Across studies the monitoring instruments were used to assess patients at baseline and at multiple time point during different modalities (i.e. HFNO, CPAP and BiPAP) and interfaces (i.e. mask and helmet) of non-invasive respiratory support. Each monitoring instrument was used to acquire multiple measurements during different modalities of non-invasive respiratory support and/or at different time points. Table 1 summarises what each advance monitoring instrument measures and how these were used across all the 78 included studies which are fully detailed in Table S1 of the supplementary materials.

Physiological responses to non-invasive respiratory support (NRS) varied according to modality, interface, pressure or flow settings, and body position, and were captured using oesophageal manometry, electrical impedance tomography (EIT), electrical activity of the diaphragm (EAdi), surface electromyography (EMG), and ultrasound. These are summarised in Table 2.

### Oesophageal manometry

Twenty-five studies (17–19,28,29,48,49,52–54,56,60–62,67,69,70,72,74,77,102,106,110,111) used oesophageal manometry to acquire measurement of oesophageal pressure (Pes) and transpulmonary driving pressure swings to quantify inspiratory and expiratory effort and the elastic load of the lung. Definition and reporting of transpulmonary pressures varied across studies, some reported dynamic values, others as quasi-static, and some omitted it; therefore, data synthesis was not feasible. Across studies, high-flow nasal oxygen (HFNO) consistently reduced inspiratory effort compared with standard oxygen therapy, with lower oesophageal pressure swings (ΔPes) observed at flow rates between 30 and 60 L/min. Increasing HFNO flow was associated with progressive reductions in ΔPes, alongside increases in end-expiratory lung volume and reductions in indices of respiratory drive. Transition from HFNO to continuous positive airway pressure (CPAP) did not produce consistent changes in ΔPes, whereas bilevel positive airway pressure (BiPAP) resulted in the greatest unloading of respiratory effort, with significantly lower ΔPes compared with baseline, HFNO, and CPAP. Notably, prone positioning during CPAP increased lung aeration but was accompanied by higher inspiratory effort (See Table 2).

### Electrical impedance tomography (EIT)

EIT was used to assess global and regional ventilation including compliance, homogeneity, ventilation distribution, and lung volume using different variables such as end-expiratory lung impedance (EELI), end-expiratory lung volume (EELV) or delta end expiratory lung impedance (ΔEELI) reported in arbitrary units (AU), millilitres (ml), or percentages (%) with no consistency across studies precluding meta-synthesis.

EIT studies demonstrated improved lung aeration when switching from face-mask oxygen to HFNO, with increases in end-expiratory lung impedance (EELI) and end-expiratory lung volume that were flow-dependent and more pronounced in dependent lung regions. Higher HFNO flows were also associated with reduced ventilation heterogeneity. Compared with HFNO, CPAP and helmet-based BiPAP produced larger increases in EELI, although effects were not uniform across interfaces and settings. During awake prone positioning, EIT-derived measures showed heterogeneous responses, with some studies reporting increased aeration and others showing redistribution of ventilation without consistent changes in global inhomogeneity indices (See Table 2).

### Electrical activity of the diaphragm (EAdi)

Electrical activity of the diaphragm (Eadi) catheter was used to measure diaphragmatic activation, neural timing and respiratory cycle to detect patient’s ventilator asynchronies and ineffective breathing pattern (42,44,45,50,55,58,64,68,90,95,104,107–109).

EAdi monitoring demonstrated improved patient–ventilator interaction when neurally adjusted ventilatory assist (NAVA) was used instead of conventional pressure support ventilation. NAVA-NIV significantly reduced the overall asynchrony index and the prevalence of severe asynchrony, shortened inspiratory trigger delay, and reduced auto-triggering, although double-triggering occurred slightly more frequently. Ineffective efforts did not differ significantly between modes as detailed in Table 2.

### Surface electromyography (EMG)

Studies using surface parasternal and diaphragmatic EMG evaluated neural respiratory drive, compensatory accessory muscle recruitment and patient’s ventilator asynchronies during BiPAP (30,47,71,75,76).

Surface parasternal EMG showed a high prevalence of patient–ventilator asynchrony during BiPAP, with more than half of patients exhibiting asynchronous events.

Severe asynchrony was associated with higher pressure support levels and greater mask leak. EMG activity generally decreased during effective BiPAP support, while higher baseline parasternal EMG activity correlated with worse lung function and predicted clinical deterioration in patients with acute COPD exacerbations (see Table 2).

### Ultrasound (US)

Respiratory muscle US was used to derive diaphragmatic thickness and thickening fraction and diaphragmatic excursion to evaluate muscle function and mechanical efficiency across patients succeeding and failing treatments (28,46,57,59,81,82,85–87,89,91,97,99,100,105,112,111).

Ultrasound studies showed that parasternal intercostal thickening fraction (PIC-TF%) and diaphragmatic thickening fraction (DTF%) were associated with non-invasive ventilation failure, with moderate-to-high discriminative performance for predicting BiPAP failure and in-hospital mortality. Lung ultrasound scores (LUS) decreased following initiation of HFNO, reflecting improved aeration over time. Diaphragmatic excursion and thickening generally decreased during BiPAP and CPAP, indicating reduced diaphragmatic contribution to ventilation during assisted breathing (Table 2).

#### 3. Detecting treatment failure

Physiological measurements of the respiratory load/capacity balance assessed by oesophageal manometry, ultrasound and electromyography were able to discriminate between patients responding and failing to non-invasive respiratory support. Specifically, patients who failed NRS had significantly higher ΔPes [MD 12.60cmH_2_O (95% CI 4.03;21.16), p=0.004], LUS score [MD 3.93 (95% CI 1.29,6.570), p=0.003] and parasternal intercostal thickening fraction (PIC-TF%) [MD 12.58% (95% CI 8.02,17.13), p<0.001] but lower diaphragmatic thickening fraction (DTF%) [MD −17.20% (95% CI −20.97,-13.42); p<0.001] and lower diaphragmatic excursion (DE) [MD - 0.95cm (95% CI −1.08,-0.82); p<0.001]. Refer to Figure 2 panel A, B, C, D and E respectively.

**Figure 2.**
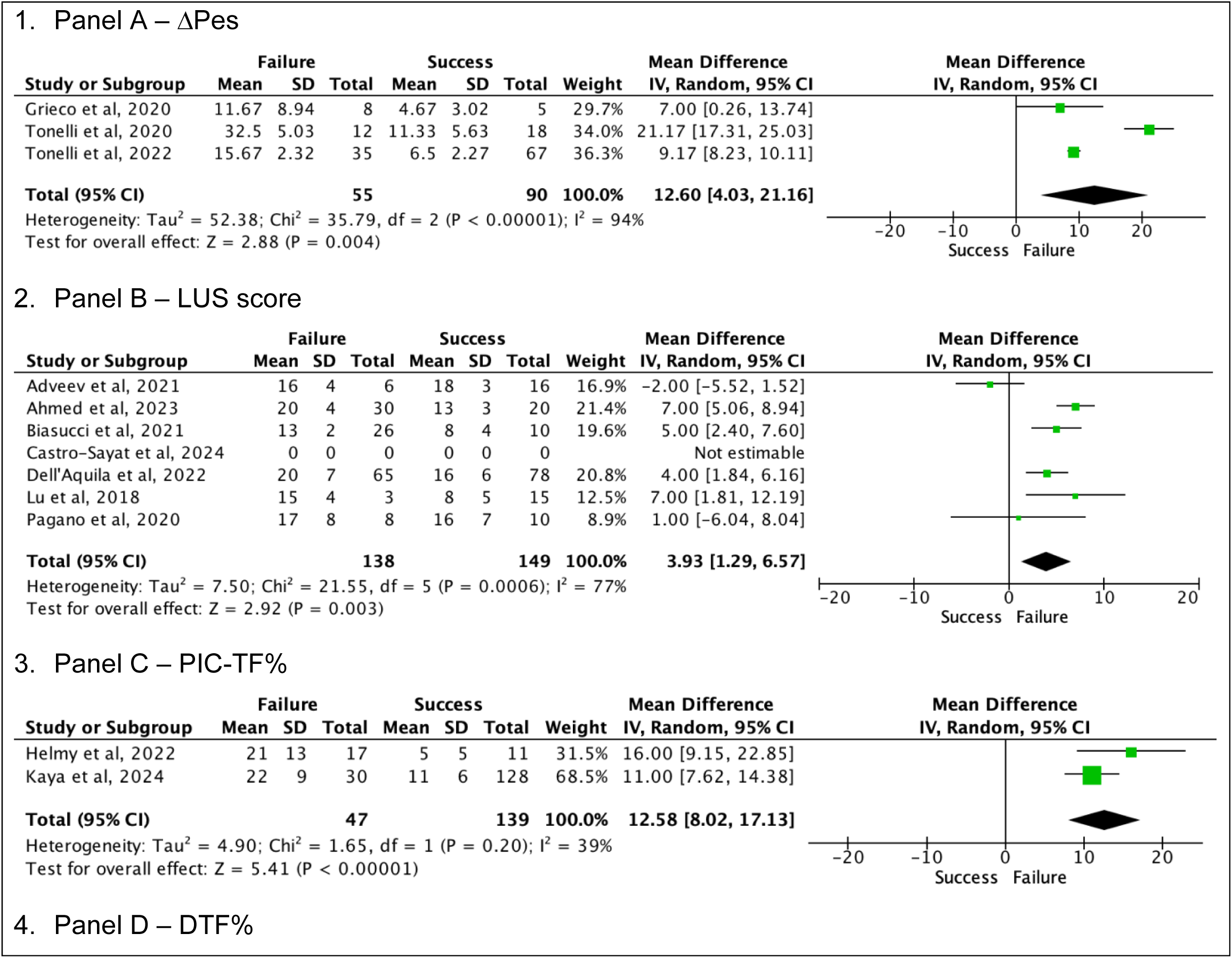
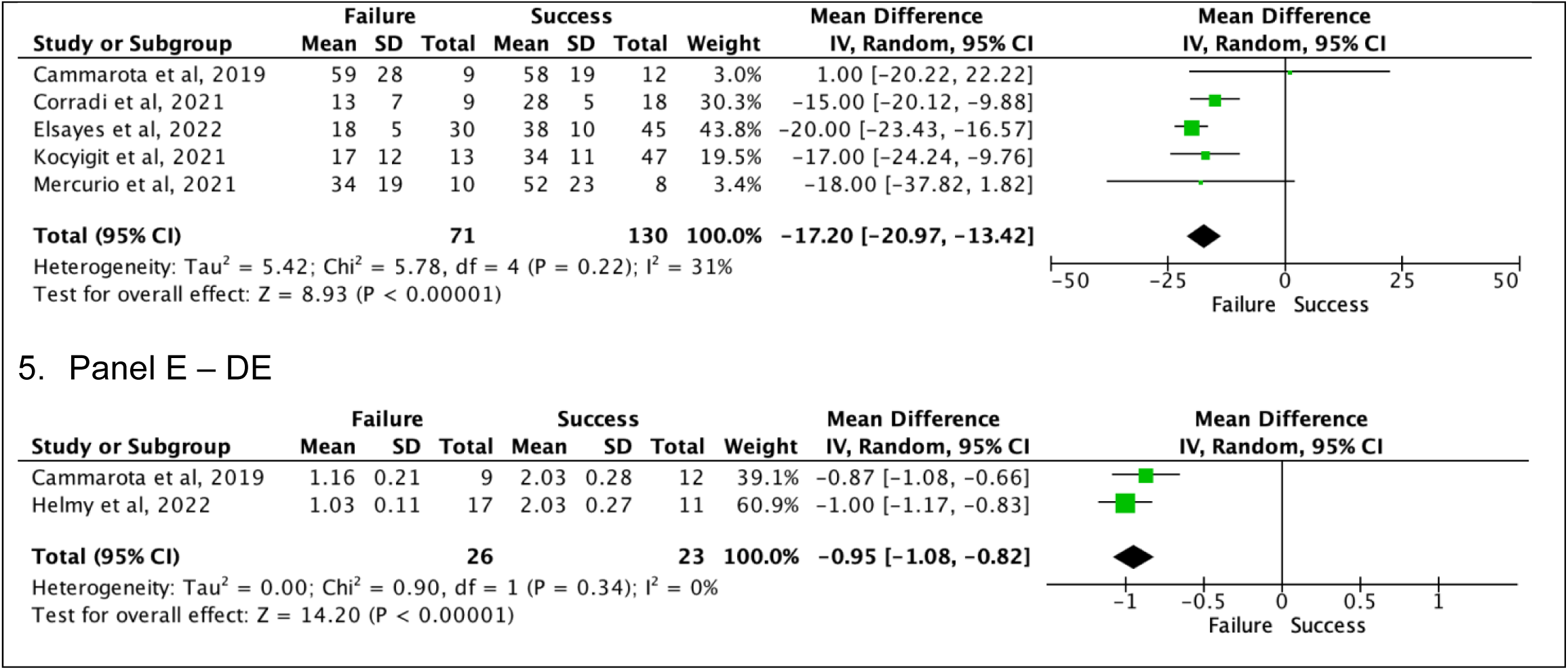
Changes in respiratory muscle workload and lung aeration between treatment failure and success of non-invasive respiratory supports. Panel A ΔPes: Delta oesophageal pressure. Panel B LUS score: Lung Ultrasound Score. Panel C PIC-TF%: parasternal intercostal thickening fraction. Panel D DTF%: diaphragmatic thickening fraction. Panel E DE: diaphragmatic excursion.

Patients with baseline ΔPes between 10 and 20 cmH_2_O were those who experienced an improvement while patients with baseline ΔPes above 30 cmH_2_O did not show any significant change in physiological nor in clinical features (18,29). All patients who did not improve showed ΔPes values above 10 cmH_2_O (18,29,53) after 2 hours on HFNO (18) and ΔPes above 30 cmH_2_O after 2 hours on facemask BiPAP (29). Patients who failed helmet NIV had ΔPes values above 10 cmH_2_O during treatment (53).

Across the five studies that used ultrasound to measure diaphragmatic excursion (DE) (82,85,86,91,103); two studies found that DE < 1.5cm was associated with BiPAP failure (82,91) and two study found that DE < 1.6cm was associated with HFNO failure (85,103).

Four studies included results of rate of intubation and hospital mortality showing no difference in intubation (odds ratio 1.15 [95% CI 0.71–1.87], P= 0.57) nor hospital mortality (odds ratio 1.12 [95% CI 0.68–1.85], P= 0.65) between NAVA-NIV and NIV alone (90,104,108,107). One study also showed no difference in 90-day mortality (22.2% vs 17.5%, p= 0.67) (104).

A study showed that surface parasternal EMG (ΔEMGpara%max) was associated with patients readmission to hospital at 14 days or death (OR 1.12, 95% 1.03 to 1.21) (30).

In two studies using EIT, patients with successful HFNO treatment had central ventilation distribution (CoV> 50%) while in the failure group, ventilation was distributed slightly toward the dorsal regions (CoV<50%); but no significant difference was noted (57,78).

## Discussion

In this systematic review we synthetised data from 78 studies from international settings including 3709 acute respiratory failure patients on non-invasive respiratory support managed across different hospital settings. Our most important finding are: 1) there are five advanced monitoring instruments that have been evaluated for clinical use (EIT, surface diaphragmatic and parasternal EMG, EAdi catheter, oesophageal manometry and diaphragmatic, parasternal, and lung US). Each instrument evaluates a different physiological domain of the respiratory load/capacity balance and lung aeration; 2) across studies, non-invasive respiratory support elicited heterogeneous physiological responses in lung aeration, respiratory effort, neural drive, and respiratory muscle function depending on modalities, settings, and patient characteristics; 3) patients that fail non-invasive respiratory support they have higher oesophageal pressure swings, higher lung ultrasound (LUS) scores, higher parasternal intercostal thickening fraction, lower diaphragmatic thickening fraction and lower diaphragmatic excursion.

### Physiological effects of non-invasive respiratory support

The clinical rational for non-invasive respiratory support is to preserve spontaneous breathing and reduce the risks related to invasive ventilation, such as sedation; yet inappropriate use can cause harm (16). Advanced monitoring showed that each treatment modality has different specific physiological effects. Across studies, HFNO consistently reduced inspiratory effort compared with standard oxygen, and higher flows (30–60 L/min) produced stepwise decreases in ΔPes; however, available data did not allow identification of an optimal flow setting for reducing inspiratory effort (18,49,60,61,67). There may be some different physiology interplay as HFNO leads to higher intrinsic PEEP and end-expiratory lung volume (60,61) washing out carbon dioxide (CO2) and lowering respiratory drive and effort (62).

Collectively, physiological effects varied across non-invasive respiratory support modalities, with CPAP predominantly affecting lung aeration and BiPAP primarily reducing inspiratory effort, although these effects were not uniformly observed across studies. However, the available data were insufficient to determine consistent dose–response relationships across patients’ groups. Physiological data showed that CPAP generally increased lung volume, as reflected by higher EELI, but this did not consistently coincided with reductions in inspiratory effort, indicating that greater aeration did not always translate into improved respiratory muscle unloading (51,74,77,106), and helmet BiPAP altered diaphragmatic mechanics with variable impact (53,54,106). Several studies reported that changes in PEEP or pressure support altered lung volume and effort to differing degrees, suggesting that adjustments in pressure settings do not uniformly yield proportional physiological effects (19,53,67,70,106). Increased pressure may improve lung recruitment and compliance thereby enhancing oxygenation, while simultaneously altering diaphragm geometry through flattening, which can reduce the mechanical efficiency to generate inspiratory pressure (114). Taken together, indiscriminate application of non-invasive respiratory support settings (i.e. flow, PEEP or PS) may contribute or aggravate the patient respiratory load-capacity balance. This highlights the need for accurate monitoring to detect potential negative effects of non-invasive respiratory support (6,7,16).

### Treatment response to non-invasive respiratory support

Our meta-analysis identified five physiological variables associated with treatment failure: higher oesophageal pressure changes (ΔPes), higher lung ultrasound (LUS) scores, increased parasternal intercostal thickening fraction, reduced diaphragmatic thickening fraction and lower diaphragmatic excursion. The combination of reduced diaphragmatic thickening and excursion, together with increased parasternal intercostal activity, suggests a work shift from the diaphragm to accessory respiratory muscles during treatment failure. This may be the physiological signature underpinning why patients do not respond to NRS (23,24,27,114–116). In our analysis, US was the most frequently used monitoring tool as it is non-invasive and portable; but this instrument has important challenges. Although reduced diaphragmatic thickening fraction (DTF%) was associated with treatment failure in our meta-analysis, its physiological interpretation remains debated. Concerns remain regarding the validity of DTF% as a surrogate for inspiratory effort, and recommends caution when using it as a standalone decision-making tool (117). Despite good reliability (86,97,100) its validity is unclear with poor correlation with oesophageal pressure (28,99,100). Additionally, diaphragm ultrasound poorly correlated with oesophageal pressure at different PEEP level (p= 0.038) (99) and pressure support (100). Internal consistency for ultrasound measurements is also unclear as the coefficients of repeatability range around 7–8 % for intra- or inter-analyser repeatability and around 15–18 % for intra- or inter-observer repeatability (100). However, a translational gap remains as only a minority of studies demonstrated that advanced monitoring directly changes clinical outcomes or reduces treatment failure (16,18,19,60,62,118).

### Strength and limitations

The strengths of this review lie in its rigorous and transparent methodology. We conducted a comprehensive systematic review and meta-analysis following PRISMA guidance, with a pre-specified protocol, systematic literature search, and structured data extraction. Where appropriate, quantitative synthesis and a meta-analysis was performed. Several limitations of the current evidence base should be acknowledged when interpreting these findings. The included studies used heterogeneous monitoring instruments, measurement definitions, and analytical approaches, limiting direct comparison and quantitative synthesis across studies. Although expert recommendations on the acquisition and interpretation of advanced respiratory monitoring exist (21,25,26,119); there remains no consensus on how these physiological parameters should be standardised or applied to guide clinical decision-making.

In addition, most studies assessed physiological responses at only two time points, typically before and after initiation of non-invasive respiratory support, without longitudinal evaluation of dynamic changes or systematic tracking of support settings. This precluded assessment of dose–response relationships between specific non-invasive respiratory support settings and clinical outcomes. Furthermore, real-world care often involves transitions between modalities, for example, alternating CPAP or BiPAP with HFNO for comfort or feeding; which were rarely captured in study designs. Finally, the patient tolerability and clinical feasibility of all advanced monitoring instruments need to be further explored to understand how they can be widely applied and implemented.

### Implications for practice and future directions

Advanced monitoring provides objective information on the respiratory load–capacity balance that complements conventional bedside assessment. While these tools can identify physiological patterns associated with treatment failure, evidence that their use improves clinical outcomes or guides personalised treatment strategies remains limited. Future work should focus on developing consensus definitions and core physiological parameters across instruments, supported by standardised acquisition and reporting frameworks. Longitudinal studies are needed to characterise how physiological measurements evolve over time with different non-invasive respiratory support modalities and how these trajectories relate to outcomes. Pragmatic evaluations should assess acceptability to clinicians and patients, the feasibility of routine use, and whether monitoring-guided strategies can meaningfully influence decision-making or outcomes. Multimodal monitoring approaches may offer added insight into the respiratory load–capacity balance, complementing rather than replacing clinical judgement and basic bedside observations. Finally, implementation studies and training programmes will be essential to ensure consistent application, reproducibility, and integration into clinical pathways.

## Conclusion

This review synthesises current evidence on advanced physiological monitoring during non-invasive respiratory support in acute respiratory failure. Our results suggest that selected physiological variables can track respiratory load–capacity balance and identify maladaptive responses to treatment. Five physiological variables: oesophageal pressure changes (i.e. ΔPes), lung ultrasound score, parasternal intercostal thickening fraction, diaphragmatic thickening fraction and diaphragmatic excursion; emerged as potential indicators associated with treatment failure. By offering objective and mechanistic assessment, a multimodal monitoring approach may support earlier identification of patients at risk of delayed intubation and inform tailoring of support. However, evidence that monitoring-guided strategies improve patient-centred outcomes remains limited, underscoring the need for prospective interventional studies.

## Data Availability

All data produced in the present work are contained in the manuscript

## Authors’ contributions

BF, ZP, TS, and JD conceived and designed the study.

BF, TS, FP, GM, EK, SN screened, extracted data and performed the analysis and quality assessment

BF drafted the manuscript

ZP, TS, LH and JD reviewed and edited the manuscript with high-level contents All authors read and approved the manuscript.

## References

1. Robbins AJ, Fowler AJ, Haines RW, Pearse RM, Prowle JR, Puthucheary Z. Emergency hospital admissions associated with non-communicable diseases 1998-2018 in England, Wales and Scotland: an ecological study. Clin Med (Lond). 2021;21(2):e179–e185.

2. Riviello ED, Kiviri W, Twagirumugabe T, Mueller A, Banner-Goodspeed VM, Officer L, Novack V, Mutumwinka M, Talmor DS, Fowler RA. Hospital Incidence and Outcomes of the Acute Respiratory Distress Syndrome Using the Kigali Modification of the Berlin Definition. Am J Respir Crit Care Med. 2016 Jan 1;193(1):52–9. doi: 10.1164/rccm.201503-0584OC.

3. Bellani G, Laffey JG, Pham T, Madotto F, Fan E, Brochard L, Esteban A, Gattinoni L, Bumbasirevic V, Piquilloud L, van Haren F, Larsson A, McAuley DF, Bauer PR, Arabi YM, Ranieri M, Antonelli M, Rubenfeld GD, Thompson BT, Wrigge H, Slutsky AS, Pesenti A; LUNG SAFE Investigators; ESICM Trials Group. Noninvasive Ventilation of Patients with Acute Respiratory Distress Syndrome. Insights from the LUNG SAFE Study. Am J Respir Crit Care Med. 2017 Jan 1;195(1):67–77. doi: 10.1164/rccm.201606-1306OC.

4. Rochwerg B, Brochard L, Elliott MW, Hess D, Hill NS, Nava S, Navalesi P Members Of The Steering Committee, Antonelli M, Brozek J, Conti G, Ferrer M, Guntupalli K, Jaber S, Keenan S, Mancebo J, Mehta S, Raoof S Members Of The Task Force. Official ERS/ATS clinical practice guidelines: noninvasive ventilation for acute respiratory failure. Eur Respir J. 2017 Aug 31;50(2):1602426. doi: 10.1183/13993003.02426-2016.

5. Grasselli, G., Calfee, C.S., Camporota, L. et al. ESICM guidelines on acute respiratory distress syndrome: definition, phenotyping and respiratory support strategies. Intensive Care Med 49, 727–759 (2023). 10.1007/s00134-023-07050-7.

6. The National Confidential Enquiry into Patient Outcome and Death. Inspiring Change. 2017. London. NCEPOD 2017. https://www.ncepod.org.uk/2017report2/downloads/InspiringChange_FullReport.pdf.

7. Independent report by the Healthcare Safety Investigation Branch. Treating COVID-19 patients using continuous positive airway pressure (CPAP) outside of a critical care unit. NHSIB Independent report by the Healthcare Safety Investigation Branch NI-003087. Available at: https://www.hsib.org.uk/investigations-and-reports/treating-covid-19-patients-using-continuous-positive-airway-pressure-cpap/.

8. Roca O, Caralt B, Messika J, Samper M, Sztrymf B, Hernandez G, et al. An index combining respiratory rate and oxygenation to predict outcome of nasal high-flow therapy. Am J Respir Crit Care Med 2019;199: 1368–1376.

9. Duan J, Han X, Bai L, Zhou L, Huang S. Assessment of heart rate, acidosis, consciousness, oxygenation, and respiratory rate to predict noninvasive ventilation failure in hypoxemic patients. Intensive Care Med 2017;43:192–199.

10. Kostakis I, Smith GB, Prytherch D, Meredith P, Price C, Chauhan A; Portsmouth Academic ConsortIum For Investigating COVID-19 (PACIFIC-19). The performance of the National Early Warning Score and National Early Warning Score 2 in hospitalised patients infected by the severe acute respiratory syndrome coronavirus 2 (SARS-CoV-2). Resuscitation. 2021 Feb;159:150–157. doi: 10.1016/j.resuscitation.2020.10.039.

11. Hao S, Caputo ND, Williams B, Ahmed A, Kelly B, Dezman ZDW, et al. Utility of skin tone on pulse oximetry in critically ill patients. Crit Care Med. 2024;52(6):e528–37.

12. Loughlin PC, Sebat F, Ritz R. Respiratory rate: the forgotten vital sign—make it count! Jt Comm J Qual Patient Saf. 2018;44(8):494–9.

13. Gerry S, Birks J, Bonnici T, Watkinson PJ, Kirtley S, Collins GS. Early warning scores for detecting deterioration in adult hospitalised patients: systematic review and critical appraisal of methodology. BMJ. 2020;369:m1501.

14. Perez, J., Brandan, L. & Telias, I. Monitoring patients with acute respiratory failure during non-invasive respiratory support to minimize harm and identify treatment failure. Crit Care 29, 147 (2025). 10.1186/s13054-025-05369-9.

15. Abe T, Takagi T, Fujii T. Update on the management of acute respiratory failure using non-invasive ventilation and pulse oximetry. Crit Care. 2023 Mar 21;27(1):92. doi: 10.1186/s13054-023-04370-4.

16. Grieco DL, Menga LS, Eleuteri D, Antonelli M. Patient self-inflicted lung injury: implications for acute hypoxemic respiratory failure and ARDS patients on non-invasive support. Minerva Anestesiol. 2019 Sep;85(9):1014–1023. doi: 10.23736/S0375-9393.19.13418-9.

17. Tonelli R, Fantini R, Tabbì L, Castaniere I, Pisani L, Pellegrino MR, Della Casa G, D’Amico R, Girardis M, Nava S, Clini EM, Marchioni A. Early Inspiratory Effort Assessment by Esophageal Manometry Predicts Noninvasive Ventilation Outcome in De Novo Respiratory Failure. A Pilot Study. Am J Respir Crit Care Med. 2020 Aug 15;202(4):558–567. doi: 10.1164/rccm.201912-2512OC.

18. Tonelli R, Fantini R, Bruzzi G, Tabbì L, Cortegiani A, Crimi C, Pisani L, Moretti A, Guidotti F, Rizzato S, Puggioni D, Vermi M, Tacconi M, Bellesia G, Ragnoli B, Castaniere I, Marchioni A, Clini E. Effect of high flow nasal oxygen on inspiratory effort of patients with acute hypoxic respiratory failure and do not intubate orders. Intern Emerg Med. 2024 Mar;19(2):333–342. doi: 10.1007/s11739-023-03471-w.

19. Tonelli R, Busani S, Tabbì L, Fantini R, Castaniere I, Biagioni E, Mussini C, Girardis M, Clini E, Marchioni A. Inspiratory Effort and Lung Mechanics in Spontaneously Breathing Patients with Acute Respiratory Failure due to COVID-19: A Matched Control Study. Am J Respir Crit Care Med. 2021 Sep 15;204(6):725–728. doi: 10.1164/rccm.202104-1029LE.

20. Tonelli R, Protti A, Spinelli E, Grieco DL, Yoshida T, Jonkman AH, Akoumianaki E, Telias I, Docci M, Rodrigues A, Perez J, Piquilloud L, Beitler J, Liu L, Roca O, Pisani L, Goligher E, Carteaux G, Bellani G, Clini E, Zhou JX, Grasselli G, Jaber S, Demoule A, Talmor D, Heunks L, Brochard L, Mauri T. Assessing inspiratory drive and effort in critically ill patients at the bedside. Crit Care. 2025 Jul 31;29(1):339. doi: 10.1186/s13054-025-05526-0.

21. Mauri T, Yoshida T, Bellani G, Goligher EC, Carteaux G, Rittayamai N, et al.; PLeUral pressure working Group (PLUG—Acute Respiratory Failure section of the European So-ciety of Intensive Care Medicine). Esophageal and transpulmonary pressure in the clinical setting: meaning, usefulness and perspectives. Intensive Care Med 2016;42: 1360–1373.

22. Brochard L, Martin GS, Blanch L, Pelosi P, Belda FJ, Jubran A, Gattinoni L, Mancebo J, Ranieri VM, Richard JC, Gommers D, Vieillard-Baron A, Pesenti A, Jaber S, Stenqvist O, Vincent JL. Clinical review: Respiratory monitoring in the ICU - a consensus of 16. Crit Care. 2012 Dec 12;16(2):219. doi: 10.1186/cc11146.

23. Spinelli E, Mauri T, Beitler JR, Pesenti A, Brodie D. Respiratory drive in the acute respiratory distress syndrome: pathophysiology, monitoring, and therapeutic interventions. Intensive Care Med. 2020 Apr;46(4):606–618. doi: 10.1007/s00134-020-05942-6.

24. Jonkman AH, de Vries HJ, Heunks LMA. Physiology of the Respiratory Drive in ICU Patients: Implications for Diagnosis and Treatment. Crit Care. 2020 Mar 24;24(1):104. doi: 10.1186/s13054-020-2776-z. Erratum in: Crit Care. 2024 Mar 21;28(1):94.

25. Tuinman, P.R., Jonkman, A.H., Dres, M. et al. Respiratory muscle ultrasonography: methodology, basic and advanced principles and clinical applications in ICU and ED pa-tients—a narrative review. Intensive Care Med 46, 594–605 (2020). 10.1007/s00134-019-05892-8.

26. Scaramuzzo G, Pavlovsky B, Adler A, et al. Electrical impedance tomography monitoring in adult ICU patients: state-of-the-art, recommendations for standardized acquisition, pro-cessing, and clinical use, and future directions. Crit Care. 2024;28(1):377.

27. Doorduin J, van Hees HW, van der Hoeven JG, Heunks LM. Monitoring of the respiratory muscles in the critically ill. Am J Respir Crit Care Med. 2013;187(1):20–7.

28. Marchioni A, Castaniere I, Tonelli R, Fantini R, Fontana M, Tabbì L, Viani A, Giaroni F, Ruggieri V, Cerri S, Clini E. Ultrasound-assessed diaphragmatic impairment is a predictor of outcomes in patients with acute exacerbation of chronic obstructive pulmonary disease undergoing noninvasive ventilation. Crit Care. 2018 Apr 27;22(1):109. doi: 10.1186/s13054-018-2033-x.

29. Tonelli R, Cortegiani A, Fantini R, Tabbì L, Castaniere I, Bruzzi G, Busani S, Ball L, Clini E, Marchioni A. Accuracy of Nasal Pressure Swing to Predict Failure of High-Flow Nasal Oxygen in Patients with Acute Hypoxemic Respiratory Failure. Am J Respir Crit Care Med. 2023 Mar 15;207(6):787–789. doi: 10.1164/rccm.202210-1848LE.

30. Suh ES, Mandal S, Harding R, Ramsay M, Kamalanathan M, Henderson K, O’Kane K, Douiri A, Hopkinson NS, Polkey MI, Rafferty G, Murphy PB, Moxham J, Hart N. Neural respiratory drive predicts clinical deterioration and safe discharge in exacerbations of COPD. Thorax. 2015 Dec;70(12):1123–30. doi: 10.1136/thoraxjnl-2015-207188.

31. Moher D, Shamseer L, Clarke M, Ghersi D, Liberati A, Petticrew M, et al. Preferred reporting items for systematic review and meta-analysis protocols (PRISMA-P) 2015 statement. Systematic Reviews. 2015;4(1):1.10.1186/2046-4053-4-1.

32. Takwoingi Y DN, Schiller I, Rücker G, Jones HE, Partlett C, Macaskill P. Chapter 10: undertaking meta-analysis. Draft version (4 October 2022) for inclusion. In: Deeks JJ, Bossuyt PM, Leeflang MM, Takwoingi Y, editors. Cochrane Handbook for Systematic Reviews of Diagnostic Test Accuracy Version 2. London: Cochrane. 2022.

33. Grieco DL, Maggiore SM, Roca O, Spinelli E, Patel BK, Thille AW, Barbas CSV, de Acilu MG, Cutuli SL, Bongiovanni F, Amato M, Frat JP, Mauri T, Kress JP, Mancebo J, Antonelli M. Non-invasive ventilatory support and high-flow nasal oxygen as first-line treatment of acute hypoxemic respiratory failure and ARDS. Intensive Care Med. 2021 Aug;47(8):851–866. doi: 10.1007/s00134-021-06459-2.

34. Dechartres A, Trinquart L, Boutron I, Ravaud P. Influence of trial sample size on treatment effect estimates: meta-epidemiological study. BMJ. 2013; 346 :f2304 doi:10.1136/bmj.f2304.

35. Morgan CJ. Use of proper statistical techniques for research studies with small samples. Am J Physiol Lung Cell Mol Physiol. 2017 Nov 1;313(5):L873–L877. doi: 10.1152/ajplung.00238.2017.

36. Delclaux C, L’Her E, Alberti C, et al. Treatment of Acute Hypoxemic Nonhypercapnic Respiratory Insufficiency With Continuous Positive Airway Pressure Delivered by a Face Mask: A Randomized Controlled Trial. JAMA. 2000;284(18):2352–2360. doi:10.1001/jama.284.18.2352.

37. Stang A. Critical evaluation of the Newcastle-Ottawa scale for the assessment of the quality of nonrandomized studies in meta-analyses. Eur J Epidemiol. 2010 Sep;25(9):603–5. doi: 10.1007/s10654-010-9491-z.

38. Higgins JPT, Thomas J, Chandler J, Cumpston M, Li T, Page MJ, Welch VA (editors). Cochrane Handbook for Systematic Reviews of Interventions version 6.3 (updated February 2022). Cochrane, 2022. Available from www.training.cochrane.org/handbook.

39. Takwoingi Y DN, Schiller I, Rücker G, Jones HE, Partlett C, Macaskill P. Supplement 1 to chapter 10: code for undertaking meta-analysis. Draft version (4 October 2022) for inclusion. In: Deeks JJ, Bossuyt PM, Leeflang MM, Takwoingi Y, editors. Cochrane handbook for systematic reviews of diagnostic test accuracy version 2. London: Cochrane. Available from www.training.cochrane.org/handbook-diagnostic-test-accuracy. 2022.

40. Deeks JJ HJ, Altman DG (editors). Chapter 10: Analysing data and undertaking meta-analyses. In: Higgins JPT, Thomas J, Chandler J, Cumpston M, Li T, Page MJ, Welch VA (editors). Cochrane Handbook for Systematic Reviews of Interventions version 6.3 (updated February 2022). Cochrane. 2022.

41. Basile, M.C., Mauri, T., Spinelli, E. et al. Nasal high flow higher than 60 L/min in patients with acute hypoxemic respiratory failure: a physiological study. Crit Care 24, 654 (2020). 10.1186/s13054-020-03344-0.

42. Bertrand PM, Futier E, Coisel Y, Matecki S, Jaber S, Constantin JM. Neurally adjusted ventilatory assist vs pressure support ventilation for noninvasive ventilation during acute respiratory failure: a crossover physiologic study. Chest. 2013 Jan;143(1):30–36. doi: 10.1378/chest.12-0424.

43. Brunelle T, Prud’homme E, Alphonsine J-E, et al. Awake prone position in COVID□19 acute respiratory failure: a randomised crossover study using electrical impedance tomography. ERJ Open Res 2023; 9: 00509–2022 [DOI: 10.1183/23120541.00509-2022].

44. Cammarota, G., Olivieri, C., Costa, R. et al. Noninvasive ventilation through a helmet in postextubation hypoxemic patients: physiologic comparison between neurally adjusted ventilatory assist and pressure support ventilation. Intensive Care Med 37, 1943–1950 (2011). 10.1007/s00134-011-2382-2.

45. Cammarota G, Longhini F, Perucca R, Ronco C, Colombo D, Messina A, Vaschetto R, Navalesi P. New Setting of Neurally Adjusted Ventilatory Assist during Noninvasive Ventilation through a Helmet. Anesthesiology. 2016 Dec;125(6):1181–1189. doi: 10.1097/ALN.0000000000001354.

46. Cammarota G, Rossi E, Vitali L, Simonte R, Sannipoli T, Anniciello F, Vetrugno L, Bignami E, Becattini C, Tesoro S, Azzolina D, Giacomucci A, Navalesi P, De Robertis E. Effect of awake prone position on diaphragmatic thickening fraction in patients assisted by noninvasive ventilation for hypoxemic acute respiratory failure related to novel coronavirus disease. Crit Care. 2021 Aug 24;25(1):305. doi: 10.1186/s13054-021-03735-x.

47. Carteaux G, Lyazidi A, Cordoba-Izquierdo A, Vignaux L, Jolliet P, Thille AW, Richard JM, Brochard L. Patient-ventilator asynchrony during noninvasive ventilation: a bench and clinical study. Chest. 2012 Aug;142(2):367–376. doi: 10.1378/chest.11-2279.

48. Ceriana P, Vitacca M, Carlucci A, Paneroni M, Pisani L, Nava S. Changes of Respiratory Mechanics in COPD Patients from Stable State to Acute Exacerbations with Respiratory Failure. COPD. 2017 Apr;14(2):150–155. doi: 10.1080/15412555.2016.1254173.

49. Delorme M, Bouchard PA, Simon M, Simard S, Lellouche F. Effects of High-Flow Nasal Cannula on the Work of Breathing in Patients Recovering From Acute Respiratory Failure. Crit Care Med. 2017 Dec;45(12):1981–1988. doi: 10.1097/CCM.0000000000002693.

50. Doorduin J, Sinderby CA, Beck J, van der Hoeven JG, Heunks LM. Automated patient-ventilator interaction analysis during neurally adjusted non-invasive ventilation and pressure support ventilation in chronic obstructive pulmonary disease. Crit Care. 2014 Oct 13;18(5):550. doi: 10.1186/s13054-014-0550-9.

51. Fossali T, Locatelli M, Colombo R, Veronese A, Borghi B, Ballone E, Castelli A, Rech R, Catena E, Ottolina D. Awake pronation with helmet CPAP in early COVID-19 ARDS patients: effects on respiratory effort and distribution of ventilation assessed by EIT. Intern Emerg Med. 2024 Oct;19(7):2025–2034. doi: 10.1007/s11739-024-03572-0.

52. Giosa, L., Collins, P.D., Sciolla, M. et al. Effects of CPAP and FiO2 on respiratory effort and lung stress in early COVID-19 pneumonia: a randomized, crossover study. Ann. Intensive Care 13, 103 (2023). 10.1186/s13613-023-01202-0.

53. Grieco DL, Menga LS, Raggi V, Bongiovanni F, Anzellotti GM, Tanzarella ES, Bocci MG, Mercurio G, Dell’Anna AM, Eleuteri D, Bello G, Maviglia R, Conti G, Maggiore SM, Antonelli M. Physiological Comparison of High-Flow Nasal Cannula and Helmet Noninvasive Ventilation in Acute Hypoxemic Respiratory Failure. Am J Respir Crit Care Med. 2020 Feb 1;201(3):303–312. doi: 10.1164/rccm.201904-0841OC.

54. Grieco, D.L., Delle Cese, L., Menga, L.S. et al. Physiological effects of awake prone position in acute hypoxemic respiratory failure. Crit Care 27, 315 (2023). 10.1186/s13054-023-04600-9.

55. Harnisch LO, Olgemoeller U, Mann J, Quintel M, Moerer O. Noninvasive Neurally Adjusted Ventilator Assist Ventilation in the Postoperative Period Produces Better Patient-Ventilator Synchrony but Not Comfort. Pulm Med. 2020 Jun 20;2020:4705042. doi: 10.1155/2020/4705042.

56. L’Her E, Deye N, Lellouche F, Taille S, Demoule A, Fraticelli A, Mancebo J, Brochard L. Physiologic effects of noninvasive ventilation during acute lung injury. Am J Respir Crit Care Med. 2005 Nov 1;172(9):1112–8. doi: 10.1164/rccm.200402-226OC.

57. Li Z, Zhang Z, Xia Q, Xu D, Qin S, Dai M, Fu F, Gao Y, Zhao Z. First Attempt at Using Electrical Impedance Tomography to Predict High Flow Nasal Cannula Therapy Outcomes at an Early Phase. Front Med (Lausanne). 2021 Oct 8;8:737810. doi: 10.3389/fmed.2021.737810.

58. Longhini F, Pan C, Xie J, Cammarota G, Bruni A, Garofalo E, Yang Y, Navalesi P, Qiu H. New setting of neurally adjusted ventilatory assist for noninvasive ventilation by facial mask: a physiologic study. Crit Care. 2017 Jul 7;21(1):170. doi: 10.1186/s13054-017-1761-7.

59. Longhini F, Pisani L, Lungu R, Comellini V, Bruni A, Garofalo E, Laura Vega M, Cammarota G, Nava S, Navalesi P. High-Flow Oxygen Therapy After Noninvasive Ventilation Interruption in Patients Recovering From Hypercapnic Acute Respiratory Failure: A Physiological Crossover Trial. Crit Care Med. 2019 Jun;47(6):e506–e511. doi: 10.1097/CCM.0000000000003740.

60. Mauri T, Alban L, Turrini C, Cambiaghi B, Carlesso E, Taccone P, Bottino N, Lissoni A, Spadaro S, Volta CA, Gattinoni L, Pesenti A, Grasselli G. Optimum support by high-flow nasal cannula in acute hypoxemic respiratory failure: effects of increasing flow rates. Intensive Care Med. 2017 Oct;43(10):1453–1463. doi: 10.1007/s00134-017-4890-1.

61. Mauri T, Turrini C, Eronia N, Grasselli G, Volta CA, Bellani G, Pesenti A. Physiologic Effects of High-Flow Nasal Cannula in Acute Hypoxemic Respiratory Failure. Am J Respir Crit Care Med. 2017 May 1;195(9):1207–1215. doi: 10.1164/rccm.201605-0916OC.

62. Mauri T, Spinelli E, Pavlovsky B, Grieco DL, Ottaviani I, Basile MC, Dalla Corte F, Pintaudi G, Garofalo E, Rundo A, Volta CA, Pesenti A, Spadaro S. Respiratory Drive in Patients with Sepsis and Septic Shock: Modulation by High-flow Nasal Cannula. Anesthesiology. 2021 Dec 1;135(6):1066–1075. doi: 10.1097/ALN.0000000000004010.

63. Öner Ö, Ergan B, Kizil AS, Gurkok MC, Dugral E, Gökmen N. Investigation of high flow nasal cannule efficiency with electric impedance tomography based parameters in COVID-19 adults patients: a retrospective study. PeerJ. 2023 Jul 14;11:e15555. doi: 10.7717/peerj.15555.

64. Piquilloud, L., Tassaux, D., Bialais, E. et al. Neurally adjusted ventilatory assist (NAVA) improves patient–ventilator interaction during non-invasive ventilation delivered by face mask. Intensive Care Med 38, 1624–1631 (2012). 10.1007/s00134-012-2626-9.

65. Rauseo M, Mirabella L, Laforgia D, Lamanna A, Vetuschi P, Soriano E, Ugliola D, Casiello E, Tullo L, Cinnella G. A Pilot Study on Electrical Impedance Tomography During CPAP Trial in Patients With Severe Acute Respiratory Syndrome Coronavirus 2 Pneumonia: The Bright Side of Non-invasive Ventilation. Front Physiol. 2021 Sep 9;12:728243. doi: 10.3389/fphys.2021.728243.

66. Rosen J, Frykholm P, Jonsson Fagerlund M, Pellegrini M, Campoccia Jalde F, von Oelreich E, et al. (2024) Lung impedance changes during awake prone positioning in COVID-19. A non□randomized cross-over study. PLoS ONE 19(2): e0299199. 10.1371/journal.pone.0299199.

67. Schifino G, Vega ML, Pisani L, Prediletto I, Catalanotti V, Comellini V, Bassi I, Zompatori M, Ranieri MV, Nava S. Effects of non-invasive respiratory supports on inspiratory effort in moderate-severe COVID-19 patients. A randomized physiological study. Eur J Intern Med. 2022 Jun;100:110–118. doi: 10.1016/j.ejim.2022.04.012.

68. Schmidt M, Dres M, Raux M, Deslandes-Boutmy E, Kindler F, Mayaux J, Similowski T, Demoule A. Neurally adjusted ventilatory assist improves patient-ventilator interaction during postextubation prophylactic noninvasive ventilation. Crit Care Med. 2012 Jun;40(6):1738–44. doi: 10.1097/CCM.0b013e3182451f77.

69. Slobod, D., Spinelli, E., Crotti, S. et al. Effects of an asymmetrical high flow nasal cannula interface in hypoxemic patients. Crit Care 27, 145 (2023). 10.1186/s13054-023-04441-6.

70. Steriade AT, Gologanu M, Bumbacea RS, Bogdan SN, Bumbacea D. Esophageal Pressure Measurement in Acute Hypercapnic Respiratory Failure Due to Severe COPD Exacerbation Requiring NIV-A Pilot Safety Study. J Clin Med. 2022 Nov 17;11(22):6810. doi: 10.3390/jcm11226810.

71. Thys F, Roeseler J, Reynaert M, Liistro G, Rodenstein DO. Noninvasive ventilation for acute respiratory failure: a prospective randomised placebo-controlled trial. Eur Respir J. 2002 Sep;20(3):545–55. doi: 10.1183/09031936.02.00287402.

72. Tonelli R, Cortegiani A, Marchioni A, Fantini R, Tabbì L, Castaniere I, Biagioni E, Busani S, Nani C, Cerbone C, Vermi M, Gozzi F, Bruzzi G, Manicardi L, Pellegrino MR, Beghè B, Girardis M, Pelosi P, Gregoretti C, Ball L, Clini E. Nasal pressure swings as the measure of inspiratory effort in spontaneously breathing patients with de novo acute respiratory failure. Crit Care. 2022 Mar 24;26(1):70. doi: 10.1186/s13054-022-03938-w.

73. Tuffet, S., Boujelben, M.A., Haudebourg, AF. et al. High flow nasal cannula and low level continuous positive airway pressure have different physiological effects during de novo acute hypoxemic respiratory failure. Ann. Intensive Care 14, 171 (2024). 10.1186/s13613-024-01408-w.

74. Vargas F, Saint-Leger M, Boyer A, Bui NH, Hilbert G. Physiologic Effects of High-Flow Nasal Cannula Oxygen in Critical Care Subjects. Respir Care. 2015 Oct;60(10):1369–76. doi: 10.4187/respcare.03814.

75. Vignaux L, Tassaux D, Carteaux G, Roeseler J, Piquilloud L, Brochard L, Jolliet P. Performance of noninvasive ventilation algorithms on ICU ventilators during pressure support: a clinical study. Intensive Care Med. 2010 Dec;36(12):2053–9. doi: 10.1007/s00134-010-1994-2.

76. Vignaux L, Vargas F, Roeseler J, Tassaux D, Thille AW, Kossowsky MP, Brochard L, Jolliet P. Patient-ventilator asynchrony during non-invasive ventilation for acute respiratory failure: a multicenter study. Intensive Care Med. 2009 May;35(5):840–6. doi: 10.1007/s00134-009-1416-5.

77. Wysocki M, Richard JC, Meshaka P. Noninvasive proportional assist ventilation compared with noninvasive pressure support ventilation in hypercapnic acute respiratory failure. Crit Care Med. 2002 Feb;30(2):323–9. doi: 10.1097/00003246-200202000-00010.

78. Zhao Z, Chang MY, Zhang T, Gow CH. Monitoring the Efficacy of High-Flow Nasal Cannula Oxygen Therapy in Patients with Acute Hypoxemic Respiratory Failure in the General Respiratory Ward: A Prospective Observational Study. Biomedicines. 2023 Nov 16;11(11):3067. doi: 10.3390/biomedicines11113067.

79. Avdeev, S.N., Nekludova, G.V., Trushenko, N.V. et al. Lung ultrasound can predict response to the prone position in awake non-intubated patients with COVID-19 associated acute respiratory distress syndrome. Crit Care 25, 35 (2021). 10.1186/s13054-021-03472-1.

80. Ahmed, Walid Mohamed Kamel MD1; Samir, Mohamed MSc2; Andraos, Ashraf Wadie MD1; Hosny, Mohamed MD1. Radiological and Clinical Perspectives to Predict Failure of Noninvasive Ventilation in Acute Respiratory Failure. The Egyptian Journal of Critical Care Medicine 10(1):p 1–6, January-March 2023. | DOI: 10.1097/EJ9.0000000000000047.

81. Antenora F, Fantini R, Iattoni A, Castaniere I, Sdanganelli A, Livrieri F, Tonelli R, Zona S, Monelli M, Clini EM, Marchioni A. Prevalence and outcomes of diaphragmatic dysfunction assessed by ultrasound technology during acute exacerbation of COPD: A pilot study. Respirology. 2017 Feb;22(2):338–344. doi: 10.1111/resp.12916.

82. Barbariol F, Deana C, Guadagnin GM, Cammarota G, Vetrugno L, Bassi F. Ultrasound diaphragmatic excursion during non-invasive ventilation in ICU: a prospective observational study. Acta Biomed. 2021 Jul 1;92(3):e2021269. doi: 10.23750/abm.v92i3.11609.

83. Biasucci DG, Buonsenso D, Piano A, Bonadia N, Vargas J, Settanni D, Bocci MG, Grieco DL, Carnicelli A, Scoppettuolo G, Eleuteri D, DE Pascale G, Pennisi MA, Franceschi F, Antonelli M; Gemelli Against COVID-19 Group. Lung ultrasound predicts non-invasive ventilation outcome in COVID-19 acute respiratory failure: a pilot study. Minerva Anestesiol. 2021 Sep;87(9):1006–1016. doi: 10.23736/S0375-9393.21.15188-0.

84. Blair PW, Siddharthan T, Liu G, Bai J, Cui E, East J, Herrera P, Anova L, Mahadevan V, Hwang J, Hossen S, Seo S, Sonuga O, Lawrence J, Peters J, Cox AL, Manabe YC, Fenstermacher K, Shea S, Rothman RE, Hansoti B, Sauer L, Crainiceanu C, Clark DV. Point-of-Care Lung Ultrasound Predicts Severe Disease and Death Due to COVID-19: A Prospective Cohort Study. Crit Care Explor. 2022 Aug 12;4(8):e0732. doi: 10.1097/CCE.0000000000000732.

85. Bruna M, Hidalgo G, Castañeda S, Galvez M, Bravo D, Benitez R, Tobar R, Quevedo J, Rodríguez J, Murua C, Madariaga R, Benavides C, Huilcaman M, Martinez F, Retamal J, Kattan E. Diaphragmatic Ultrasound Predictors of High-Flow Nasal Cannula Therapeutic Failure in Critically Ill Patients With SARS-CoV-2 Pneumonia. J Ultrasound Med. 2023 Jun;42(6):1277–1284. doi: 10.1002/jum.16141.

86. Cammarota G, Sguazzotti I, Zanoni M, Messina A, Colombo D, Vignazia GL, Vetrugno L, Garofalo E, Bruni A, Navalesi P, Avanzi GC, Della Corte F, Volpicelli G, Vaschetto R. Diaphragmatic Ultrasound Assessment in Subjects With Acute Hypercapnic Respiratory Failure Admitted to the Emergency Department. Respir Care. 2019 Dec;64(12):1469–1477. doi: 10.4187/respcare.06803.

87. Corradi F, Vetrugno L, Orso D, Bove T, Schreiber A, Boero E, Santori G, Isirdi A, Barbieri G, Forfori F. Diaphragmatic thickening fraction as a potential predictor of response to continuous positive airway pressure ventilation in Covid-19 pneumonia: A single-center pilot study. Respir Physiol Neurobiol. 2021 Feb;284:103585. doi: 10.1016/j.resp.2020.103585.

88. Dell’Aquila P, Raimondo P, Racanelli V, De Luca P, De Matteis S, Pistone A, Melodia R, Crudele L, Lomazzo D, Solimando AG, Moschetta A, Vacca A, Grasso S, Procacci V, Orso D, Vetrugno L. Integrated lung ultrasound score for early clinical decision-making in patients with COVID-19: results and implications. Ultrasound J. 2022 Jun 1;14(1):21. doi: 10.1186/s13089-022-00264-8.

89. Elsayed, A. A., Neanaa, E. H. M., & Beshey, B. N. (2022). Diaphragmatic impairment as a predictor of invasive ventilation in acute exacerbation of chronic obstructive pulmonary disease patients. Egyptian Journal of Anaesthesia, 38(1), 334–341. 10.1080/11101849.2022.2085975.

90. Hansen KK, Jensen HI, Andersen TS, Christiansen CF. Intubation rate, duration of noninvasive ventilation and mortality after noninvasive neurally adjusted ventilatory assist (NIV-NAVA). Acta Anaesthesiol Scand. 2020 Mar;64(3):309–318. doi: 10.1111/aas.13499.

91. Helmy MA, Hasanin A, Milad LM, Mostafa M, Fathy S. Parasternal intercostal muscle thickening as a predictor of non-invasive ventilation failure in patients with COVID-19. Anaesth Crit Care Pain Med. 2022 Jun;41(3):101063. doi: 10.1016/j.accpm.2022.101063.

92. Ji L, Cao C, Gao Y, Zhang W, Xie Y, Duan Y, Kong S, You M, Ma R, Jiang L, Liu J, Sun Z, Zhang Z, Wang J, Yang Y, Lv Q, Zhang L, Li Y, Zhang J, Xie M. Prognostic value of bedside lung ultrasound score in patients with COVID-19. Crit Care. 2020 Dec 22;24(1):700. doi: 10.1186/s13054-020-03416-1.

93. Kaya AG, Öz M, Erol S, Arslan F, Çiledağ A, Kaya A. Intercostal Muscle Function During Noninvasive Ventilation and Acute Hypercapnic Respiratory Failure. Respir Care. 2024 Jul 24;69(8):982–989. doi: 10.4187/respcare.11676.

94. Lichter Y, Topilsky Y, Taieb P, Banai A, Hochstadt A, Merdler I, Gal Oz A, Vine J, Goren O, Cohen B, Sapir O, Granot Y, Mann T, Friedman S, Angel Y, Adi N, Laufer-Perl M, Ingbir M, Arbel Y, Matot I, Szekely Y. Lung ultrasound predicts clinical course and outcomes in COVID-19 patients. Intensive Care Med. 2020 Oct;46(10):1873–1883. doi: 10.1007/s00134-020-06212-1.

95. Longhini F, Liu L, Pan C, Xie J, Cammarota G, Bruni A, Garofalo E, Yang Y, Navalesi P, Qiu H. Neurally-Adjusted Ventilatory Assist for Noninvasive Ventilation via a Helmet in Subjects With COPD Exacerbation: A Physiologic Study. Respir Care. 2019 May;64(5):582–589. doi: 10.4187/respcare.06502.

96. Lu X, Wu C, Gao Y, Zhang M. Bedside Ultrasound Assessment of Lung Reaeration in Patients With Blunt Thoracic Injury Receiving High-Flow Nasal Cannula Oxygen Therapy: A Retrospective Study. J Intensive Care Med. 2020 Oct;35(10):1095–1103. doi: 10.1177/0885066618815649.

97. Mercurio G, D’Arrigo S, Moroni R, Grieco DL, Menga LS, Romano A, Annetta MG, Bocci MG, Eleuteri D, Bello G, Montini L, Pennisi MA, Conti G, Antonelli M. Diaphragm thickening fraction predicts noninvasive ventilation outcome: a preliminary physiological study. Crit Care. 2021 Jun 26;25(1):219. doi: 10.1186/s13054-021-03638-x.

98. Pagano A, Porta G, Bosso G, Allegorico E, Serra C, Dello Vicario F, Minerva V, Russo T, Altruda C, Arbo P, Mercurio V, Numis FG. Non-invasive CPAP in mild and moderate ARDS secondary to SARS-CoV-2. Respir Physiol Neurobiol. 2020 Sep;280:103489. doi: 10.1016/j.resp.2020.103489.

99. Steinberg I, Chiodaroli E, Gattarello S, Cappio Borlino S, Chiumello D. Diaphragmatic ultrasound and esophageal pressure in COVID-19 pneumonia during helmet CPAP. Intensive Care Med. 2022 Aug;48(8):1095–1096. doi: 10.1007/s00134-022-06785-z.

100. Vivier E, Mekontso Dessap A, Dimassi S, Vargas F, Lyazidi A, Thille AW, Brochard L. Diaphragm ultrasonography to estimate the work of breathing during non-invasive ventilation. Intensive Care Med. 2012 May;38(5):796–803. doi: 10.1007/s00134-012-2547-7.

101. Artaud-Macari E, Bubenheim M, Le Bouar G, et al. High-flow oxygen therapy versus noninvasive ventilation: a randomised physiological crossover study of alveolar recruitment in acute respiratory failure. ERJ Open Res 2021; 7: 00373–2021 [DOI: 10.1183/23120541.00373-2021].

102. Basoalto, R., Damiani, L.F., Jalil, Y. et al. Physiological effects of high-flow nasal cannula oxygen therapy after extubation: a randomized crossover study. Ann. Intensive Care 13, 104 (2023). 10.1186/s13613-023-01203-z.

103. Boscolo, A., Pettenuzzo, T., Zarantonello, F. et al. Asymmetrical high-flow nasal cannula performs similarly to standard interface in patients with acute hypoxemic post-extubation respiratory failure: a pilot study. BMC Pulm Med 24, 21 (2024). 10.1186/s12890-023-02820-x.

104. Chhabria BA, Prasad KT, Dhooria S, Muthu V, Aggarwal AN, Agarwal R, Gandra RR, Sehgal IS. A randomized controlled trial comparing non-invasive ventilation delivered using neurally adjusted ventilator assist (NAVA) or adaptive support ventilation (ASV) in patients with acute exacerbation of chronic obstructive pulmonary disease. J Crit Care. 2023 Jun;75:154250. doi: 10.1016/j.jcrc.2022.154250.

105. Laverdure F, Genty T, Rezaiguia-Delclaux S, Herve P, Stephan F. Ultrasound Assessment of Respiratory Workload With High-Flow Nasal Oxygen Versus Other Noninvasive Methods After Chest Surgery. J Cardiothorac Vasc Anesth. 2019 Nov;33(11):3042–3047. doi: 10.1053/j.jvca.2019.05.020.

106. Menga LS, Delle Cese L, Rosà T, Cesarano M, Scarascia R, Michi T, Biasucci DG, Ruggiero E, Dell’Anna AM, Cutuli SL, Tanzarella ES, Pintaudi G, De Pascale G, Sandroni C, Maggiore SM, Grieco DL, Antonelli M. Respective Effects of Helmet Pressure Support, Continuous Positive Airway Pressure, and Nasal High-Flow in Hypoxemic Respiratory Failure: A Randomized Crossover Clinical Trial. Am J Respir Crit Care Med. 2023 May 15;207(10):1310–1323. doi: 10.1164/rccm.202204-0629OC. Erratum in: Am J Respir Crit Care Med. 2023 Nov 1;208(9):1004. doi: 10.1164/rccm.v208erratum3.

107. Prasad KT, Gandra RR, Dhooria S, Muthu V, Aggarwal AN, Agarwal R, Sehgal IS. Comparing Noninvasive Ventilation Delivered Using Neurally-Adjusted Ventilatory Assist or Pressure Support in Acute Respiratory Failure. Respir Care. 2021 Feb;66(2):213–220. doi: 10.4187/respcare.07952.

108. Tajamul S, Hadda V, Madan K, Tiwari P, Mittal S, Khan MA, Mohan A, Guleria R. Neurally-Adjusted Ventilatory Assist Versus Noninvasive Pressure Support Ventilation in COPD Exacerbation: The NAVA-NICE Trial. Respir Care. 2020 Jan;65(1):53–61. doi: 10.4187/respcare.07122.

109. Wang DQ, Luo J, Xiong XH, Zhu LH, Zhang WW. Effect of non-invasive NAVA on the patients with acute exacerbation of chronic obstructive pulmonary disease. Zhonghua Yi Xue Za Zhi. 2016 Nov 15;96(42):3375–3378. doi: 10.3760/cma.j.issn.0376-2491.2016.42.004.

110. Lassola S, Miori S, Sanna A, Cucino A, Magnoni S, Umbrello M. Central venous pressure swing outperforms diaphragm ultrasound as a measure of inspiratory effort during pressure support ventilation in COVID-19 patients. J Clin Monit Comput. 2022 Apr;36(2):461–471. doi: 10.1007/s10877-021-00674-4.

111. Vega ML., Schifino G, Pisani L, Catalanotti V, Prediletto I, Nava S Diaphragm thickening fraction and inspiratory effort in patients with SARS-COV II pneumonia receiving different non-invasive respiratory supports. Pulmonology, 2023; 29(5), 424–427. 10.1016/j.pulmoe.2023.02.001.

112. Kocyigit H, Gunalp M, Genc S, Oguz AB, Koca A, Polat O. Diaphragm dysfunction detected with ultrasound to predict noninvasive mechanical ventilation failure: A prospective cohort study. Am J Emerg Med. 2021 Jul;45:202–207. doi: 10.1016/j.ajem.2020.08.014.

113. Castro-Sayat M, Colaianni-Alfonso N, Vetrugno L, Olaizola G, Benay C, Herrera F, Saá Y, Montiel G, Haedo S, Previgliano I, Toledo A, Siroti C. Lung ultrasound score predicts outcomes in patients with acute respiratory failure secondary to COVID-19 treated with non-invasive respiratory support: a prospective cohort study. Ultrasound J. 2024 Mar 8;16(1):20. doi: 10.1186/s13089-024-00365-6.

114. Jansen D, Jonkman AH, Vries HJ, Wennen M, Elshof J, Hoofs MA, van den Berg M, Man AME, Keijzer C, Scheffer GJ, van der Hoeven JG, Girbes A, Tuinman PR, Marcus JT, Ottenheijm CAC, Heunks L. Positive end-expiratory pressure affects geometry and function of the human diaphragm. J Appl Physiol (1985). 2021 Oct 1;131(4):1328–1339. doi: 10.1152/japplphysiol.00184.2021.

115. Dres M, Goligher EC, Heunks L, Brochard LJ. Diaphragm dysfunction: diagnostic approaches and management strategies. Intensive Care Med. 2018;44(9):1441–52.

116. Sinderby C, Navalesi P, Beck J, Skrobik Y, Comtois N, Friberg S, et al. Neural control of mechanical ventilation in respiratory failure. Nat Med. 1999;5(12):1433–6.

117. Hermans, G., Demoule, A. & Heunks, L. How I perform diaphragmatic ultrasound in the intensive care unit. Intensive Care Med 50, 2175–2178 (2024). 10.1007/s00134-024-07688-x.

118. Grieco DL, Menga LS, Cesarano M, et al. Effect of Helmet Noninvasive Ventilation vs High-Flow Nasal Oxygen on Days Free of Respiratory Support in Patients With COVID-19 and Moderate to Severe Hypoxemic Respiratory Failure: The HENIVOT Randomized Clinical Trial. JAMA. 2021;325(17):1731–1743. doi:10.1001/jama.2021.4682.

119. Jonkman, A.H., Warnaar, R.S.P., Baccinelli, W. et al. Analysis and applications of respiratory surface EMG: report of a round table meeting. Crit Care 28, 2 (2024). 10.1186/s13054-023-04779-x.

